# Evaluation of Confocal Laser Endomicroscopy for Detection of Occult Gastric Carcinoma in *CDH1* Variant Carriers

**DOI:** 10.1101/2020.06.01.20118471

**Authors:** Samuel A. Schueler, Lauren A. Gamble, Bryan F. Curtin, Samantha M. Ruff, Maureen Connolly, Cathleen Hannah, Martha Quezado, Markku Miettinen, Maureen George, Andrew M. Blakely, Jonathan M. Hernandez, Theo Heller, Christopher Koh, Jeremy L. Davis

## Abstract

**Background & Aims:** Hereditary diffuse gastric cancer, attributed to inactivating germline *CDH1* variants, is characterized by signet ring cell (SRC) morphology. We sought to evaluate the occult cancer detection rate using probe-based confocal laser endomicroscopy (pCLE) during endoscopic surveillance.

**Methods:** A prospective, single-institution study was conducted in asymptomatic adults with pathogenic or likely pathogenic (P/LP) *CDH1* variants. Subjects received endoscopic gastric surveillance using pCLE in conjunction with the consensus Cambridge method (CM) of non-targeted mucosal biopsies. Systematic examination was performed with white light endoscopy (WLE) and pCLE. Abnormalities visualized by pCLE were biopsied, followed by non-targeted mucosal biopsies according to the CM. Pathologists were blinded to clinical and endomicroscopic findings. Primary endpoint was to determine pCLE sensitivity for detection of occult SRC carcinoma compared to CM.

**Results:** Thirty-six patients with P/LP *CDH1* variants underwent endoscopy using pCLE and CM. Majority were female (75%) with median age 47 years. Median procedure time was 52.5 minutes, without serious adverse events. Targeted biopsies of focal abnormalities on WLE were negative for carcinoma. Overall, 19.4% (7/36) patients had SRC detected on ≥1 biopsy. Non-targeted CM biopsies revealed SRC in 11.1% (4/36), whereas pCLE revealed SRC in 16.7% (6/36). Fifteen patients, 5 of whom had SRC at endoscopy, underwent total gastrectomy; all 15 explants contained occult carcinoma. In those 15 patients, the false-negative SRC detection rates for pCLE and CM were 67% and 87%, respectively.

**Conclusions:** Confocal endomicroscopy alone has low sensitivity for occult cancer detection in *CDH1* variant carriers. Reliable endoscopic surveillance is lacking as an alternative to prophylactic surgery in this high-risk population. (http://ClinicalTrials.gov, Number: NCT03648879).

## Introduction

Hereditary causes of gastric adenocarcinoma account for approximately 1–3% of incident cases, of which the most common is hereditary diffuse gastric cancer (HDGC).^1, 2^ HDGC is attributed to inactivating germline variants in the *CDH1* tumor suppressor gene in approximately 20–40% of cases.^2–4^ The *CDH1* gene encodes the glycoprotein E-cadherin, which is located on the surface of epithelial cells and plays a crucial role in cell-cell adhesion. Cancer cells with metastatic potential often demonstrate loss of E-cadherin expression.^5^ There are more than 100 known pathogenic germline variants in the *CDH1* gene.^2, 6^ Asymptomatic carriers of pathogenic *CDH1* variants harbor occult foci of intramucosal signet ring cell (SRC) carcinomas in the absence of gross mucosal abnormalities, starting at a young age.^4^ The lifetime risk of developing advanced diffuse-type gastric cancer (DGC) in *CDH1* variant carriers is 33–42% based on revised penetrance estimates.^7, 8^ Prophylactic total gastrectomy is recommended in variant carriers starting as early as 20 years of age.^2, 4^ However, total gastrectomy is associated with life-long morbidity that includes major weight loss and micronutrient deficiencies.^9, 10^

An alternative to prophylactic surgery is regular endoscopic surveillance of the gastric mucosa. For patients who choose to delay or forego total gastrectomy, annual high-definition white light endoscopy (WLE) with six biopsies from each anatomical zone of the stomach (antrum, transitional zone, body, fundus, and cardia) and any other visible lesions is recommended, and is referred to as the Cambridge method (CM).^4, 11^ However, endoscopic surveillance with random mucosal biopsies in these patients carries an unacceptably high (up to 96%) false-negative rate.^12 13, 14^ For reference, occult, *in situ* or early-stage (T1a) SRC carcinomas are found in up to 100% of gastrectomy explants from asymptomatic carriers of *CDH1* pathogenic variants.^15, 16^ The false negative rate of random endoscopic biopsy is attributed to the large surface area of gastric mucosa and the typical sub-epithelial location of signet ring cancer cells.

Probe-based confocal laser endomicroscopy (pCLE) enables endoscopists to obtain *in vivo* histologic images of gastrointestinal mucosa.^17, 18^ A fiberoptic probe inserted through a standard endoscope is used to image the microstructure of mucosal tissues including, but not limited to, the identification of cells and vessels and their organization or architecture. Until now, pCLE has not been evaluated as a surveillance modality in patients with HDGC due to pathogenic germline *CDH1* variants. We propose this technique may afford a more sensitive method of surveillance of occult gastric cancer in asymptomatic carriers of pathogenic *CDH1* variants. We conducted a clinical trial to determine if pCLE provides greater sensitivity for detection of SRC foci compared to the Cambridge method, and to define the false negative rate of pCLE cancer detection in patients who subsequently choose prophylactic total gastrectomy.

## Methods

### Study population

A single-arm, phase II clinical trial was approved by the institutional review board and was open to patient enrollment from February 2019 through October 2019. Eligible patients were asymptomatic carriers of pathogenic or likely pathogenic (P/LP) *CDH1* variants, 18 years or older, physiologically able to undergo upper endoscopy, and able to provide written informed consent. Patients with concurrent illness that would preclude upper gastrointestinal endoscopy were excluded; specifically, unstable angina, recent (within 3 months) myocardial infarction, contraindications to general anesthesia, and known bleeding disorders. All patients underwent high definition WLE with a 2T endoscope to facilitate pCLE and cold forceps biopsy nearly simultaneously (Olympus GIF-2TH180 endoscope, Global Endoscopy Solutions). All patients were administered monitored anesthesia care or general anesthesia. Examinations began with digital image capture under WLE from 22 areas of the stomach using a previously described systematic mapping schema.^19^ These areas include antegrade 4-section views (anterior wall, lesser curve, posterior wall, and greater curve) of the antrum, lower body, and middle upper body followed by retrograde 4-section views of the fundus and retrograde 3-section views (anterior wall, lesser curve, posterior wall) of incisura, middle upper body and cardia/fundus (Supplemental Figure 1). Probe-based CLE was performed before any biopsy of focal abnormality, but only after complete WLE examination and image capture of all twenty-two areas. Next, intravenous fluorescein (250mg; 2.5mL of 100mg/mL solution) was administered followed by 10mL saline flush. Probe-based CLE evaluation was performed immediately following fluorescein injection (Cellvizio® 100 series systems with Confocal Miniprobes^™^, Mauna Kea Technologies). Two study investigators were present and conducted real-time confocal analysis of each of the 22 aforementioned areas of the stomach by extending the probe onto the surface of normal-appearing gastric mucosa. Digital video and still image capture were performed to document both regular and irregular confocal findings from each section. Bite-on-bite gastric biopsies were obtained during the confocal exam with agreement by the investigators. Cold biopsy forceps with radial jaw and needle was used (Boston Scientific, 2.8mm). After conclusion of pCLE evaluation, non-targeted gastric biopsies were obtained according to the Cambridge method; 6 biopsies each from antrum, transitional zone, body, cardia and fundus.^11^ Abnormal findings via WLE, such as pale mucosal areas, were also recorded and/or biopsied separately during the procedure, but only after pCLE evaluation to avoid image interference from mucosal disruption. All samples were placed in formalin and labeled according to anatomic location. Images obtained by pCLE and WLE images were annotated in conjunction with tissue biopsy location data. Biopsy tissue was processed through cycles of formalin, alcohol, xylene, and paraffin blocks cut 4-microns thick, which were then stained with hematoxylin and eosin. All biopsies were examined by two experienced gastrointestinal pathologists (MM, MQ) blinded to clinical endoscopic findings and source of biopsies (e.g. Cambridge or pCLE). All biopsies and resection specimens underwent staining with hematoxylin and eosin. Periodic acid-Schiff staining was performed in all biopsies and selected sections of resection specimens to facilitate identification of signet ring cells. Biopsies revealing occult carcinomas were reviewed by the principal investigator after pathologic consensus diagnosis. All patients were notified of biopsy findings and received appropriate counseling regarding gastric cancer risk.

### Statistical analysis

Based on our institutional occult cancer detection rate of 24% with surveillance endoscopy, 36 patients undergoing pCLE would give 89% power to rule out a 24% rate of detection in favor of a 45% rate of detection with a one-sided 0.10 significance level exact binomial test. An accrual ceiling of 40 was set to allow for a small number of unevaluable patients. Standard analysis was performed on the two tests (pCLE and CM) including sensitivity with a paired-samples analysis, false negative rates, and diagnostic accuracy was estimated using 95% confidence interval. Fisher exact test was used to compare the biopsy positivity rate of pCLE and CM. All authors had access to the study data and reviewed and approved the final manuscript.

## Results

Thirty-seven patients with *CDH1* variants underwent WLE and pCLE examination per protocol. Thirty-six patients harbored a P/LP *CDH1* variant and are the subject of analysis (Table 1). One patient who underwent pCLE was later determined to carry a *CDH1* variant of uncertain significance and was removed from study. The majority (75%) of patients were female, 89% were Caucasian, median age was 47 years (range 25–74), 61% previously underwent at least one upper endoscopy. Median procedure time was 52.5 minutes. Less than half the study population (36.1%) met consensus criteria for HDGC genetic testing. Pathogenic or likely pathogenic *CDH1* variants were: 9 nonsense, 8 frameshift, 7 cryptic splice, 5 canonical splice, 4 intronic splicing, and 3 deletion (Table 1).

**Table 1:**
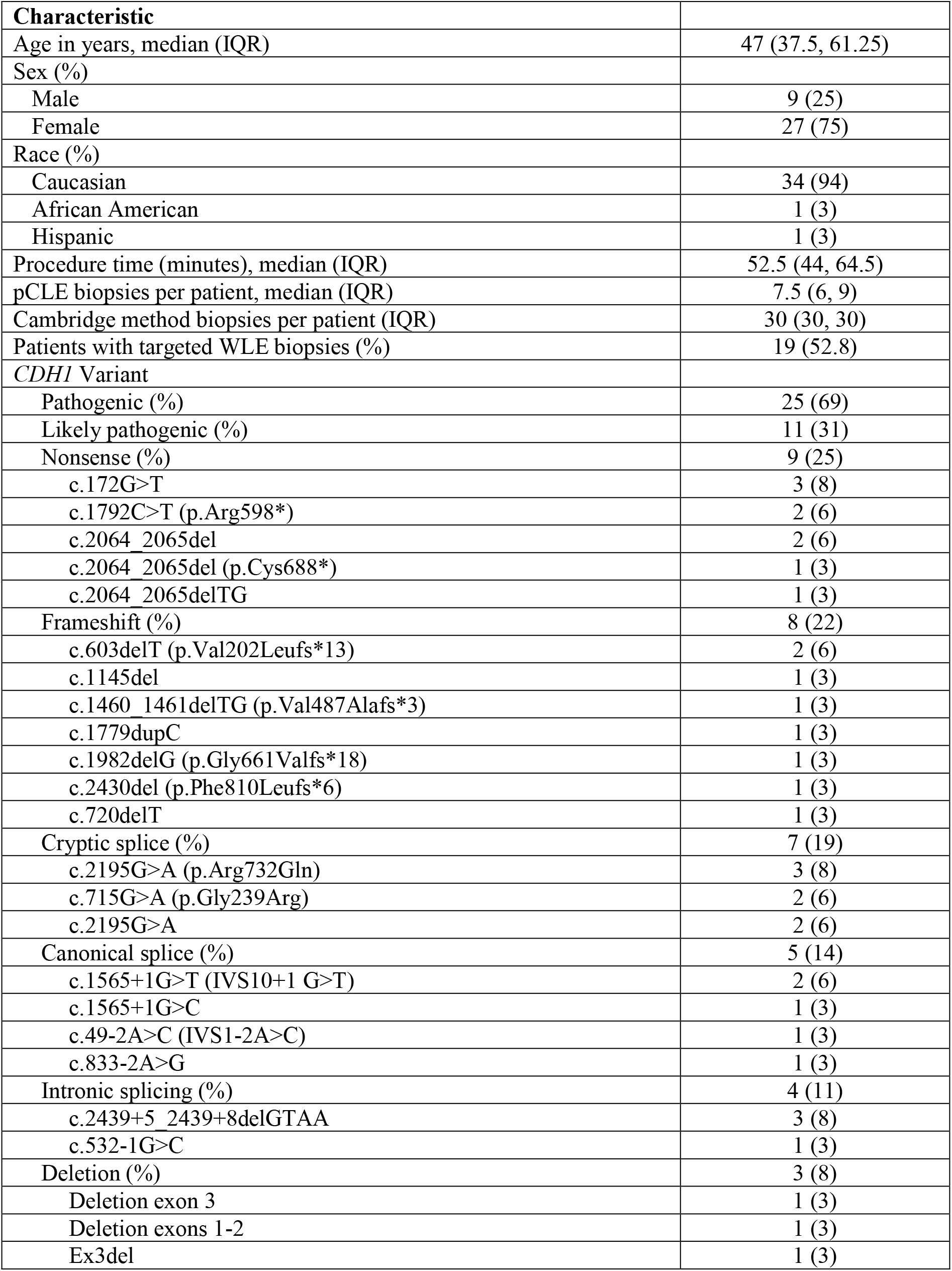
Patient demographics and pCLE procedure characteristics.

### WLE endoscopy findings

Standard WLE endoscopy identified mucosal abnormalities in 30/36 (83.3%) patients, including erythematous, erosive, or ulcerated mucosa (12/36, 33.3%), gastric polyps (11/36, 30.1%), gastric nodule(s)/papule(s) or nodular-appearing mucosa (9/36, 25%), mucosal atrophy (4/36, 11.1%), friable mucosa (4/36, 11.1%), or white spots (2/36, 5.6%). Nineteen of 36 (52.8%) patients underwent either 1 or 2 mucosal biopsies due to WLE findings. Subsequent pathologic analysis of these targeted biopsies revealed gastritis in 8/36 (22.2%) and fundic gland polyps in 4/36 (11.1%). One patient had biopsy evidence of *Helicobacter pylori* infection. WLE with targeted biopsies failed to identify occult SRC carcinoma (Supplemental Table 1). The remaining six patients (16.7%) had normal-appearing gastric mucosa on WLE examination.

### Surveillance biopsy results

Overall, 7 of 36 (19.4%) patients had occult SRC foci discovered on thirteen separate biopsies during surveillance endoscopy. Eleven of those 13 (84.6%) positive biopsies were obtained from the gastric fundus. In total, 282 pCLE-guided biopsies were obtained with a median of 7.5 per patient (IQR 6, 9). Six patients (16.7%, 6/36) underwent pCLE-directed gastric biopsy that contained occult SRC carcinoma. This consisted of nine separate pCLE biopsies, which revealed a cancer focus of median size 0.5 mm (IQR 0.25, 1.25) in the following anatomic locations: posterior fundus (4 biopsies), anterior fundus (3), posterior upper middle body (1), and retroflex view lesser upper middle body (1). Abnormal pCLE images with corresponding biopsy photomicrographs (Figure 1) illustrate the irregular appearance of displaced gastric pits, which was not specific for cancer. Typical pCLE images of gastric mucosa (Figure 2) demonstrated no pathologic changes on biopsy.

**Figure 1.**
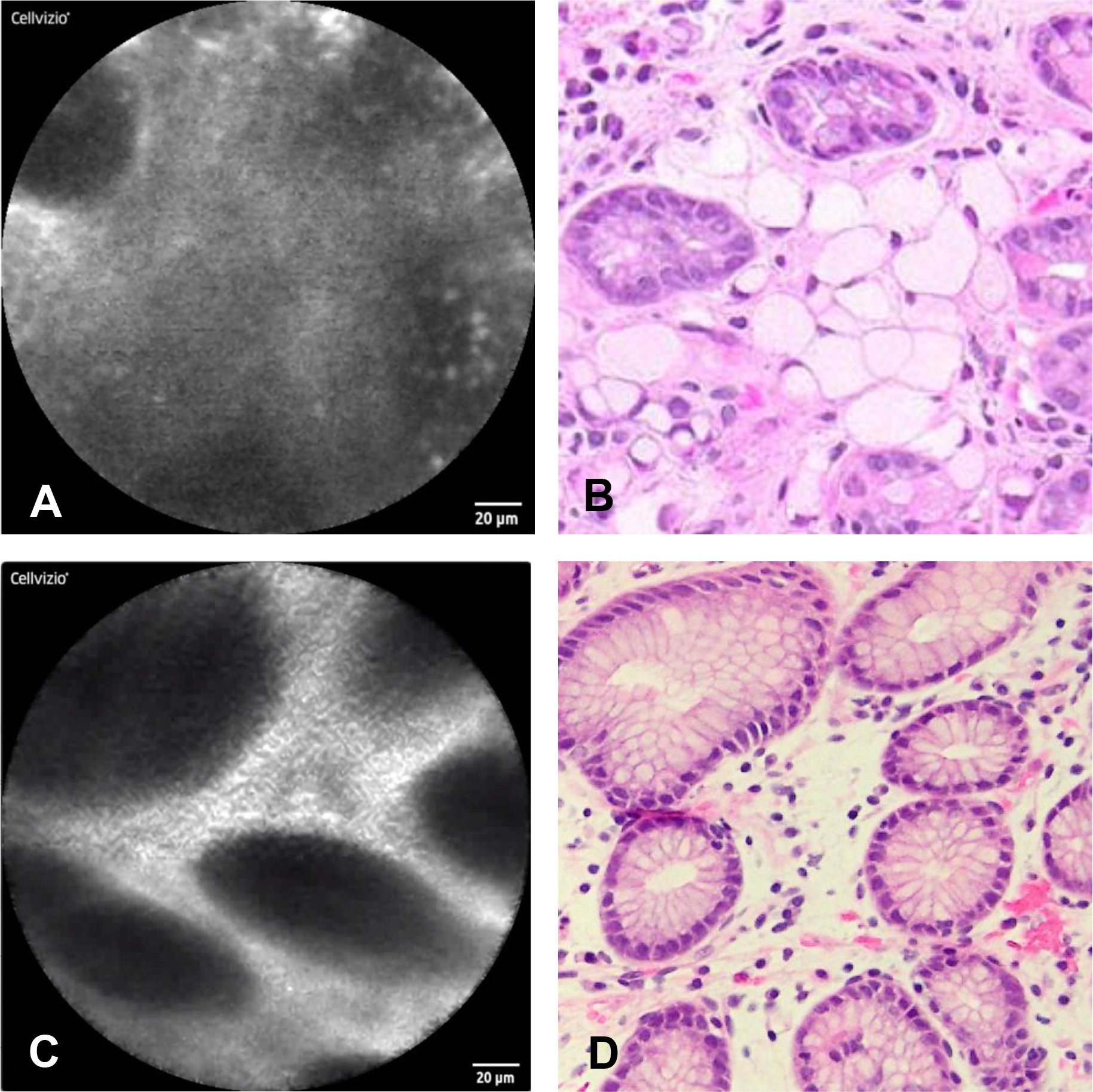
Atypical pCLE images of gastric mucosa with matched biopsy photomicrographs. Abnormal appearing confocal laser endomicroscopic image (a) with cross-sectional view of gastric pits displaced by the expansion of the lamina propria by signet ring cells and (b) corresponding biopsy (hematoxylin and eosin, 40x). Abnormal appearing confocal laser endomicroscopic image (c) with cross-sectional view of gastric pits displaced by stromal edema within the lamina propria (d) corresponding biopsy (hematoxylin and eosin, 40x).

**Figure 2.**
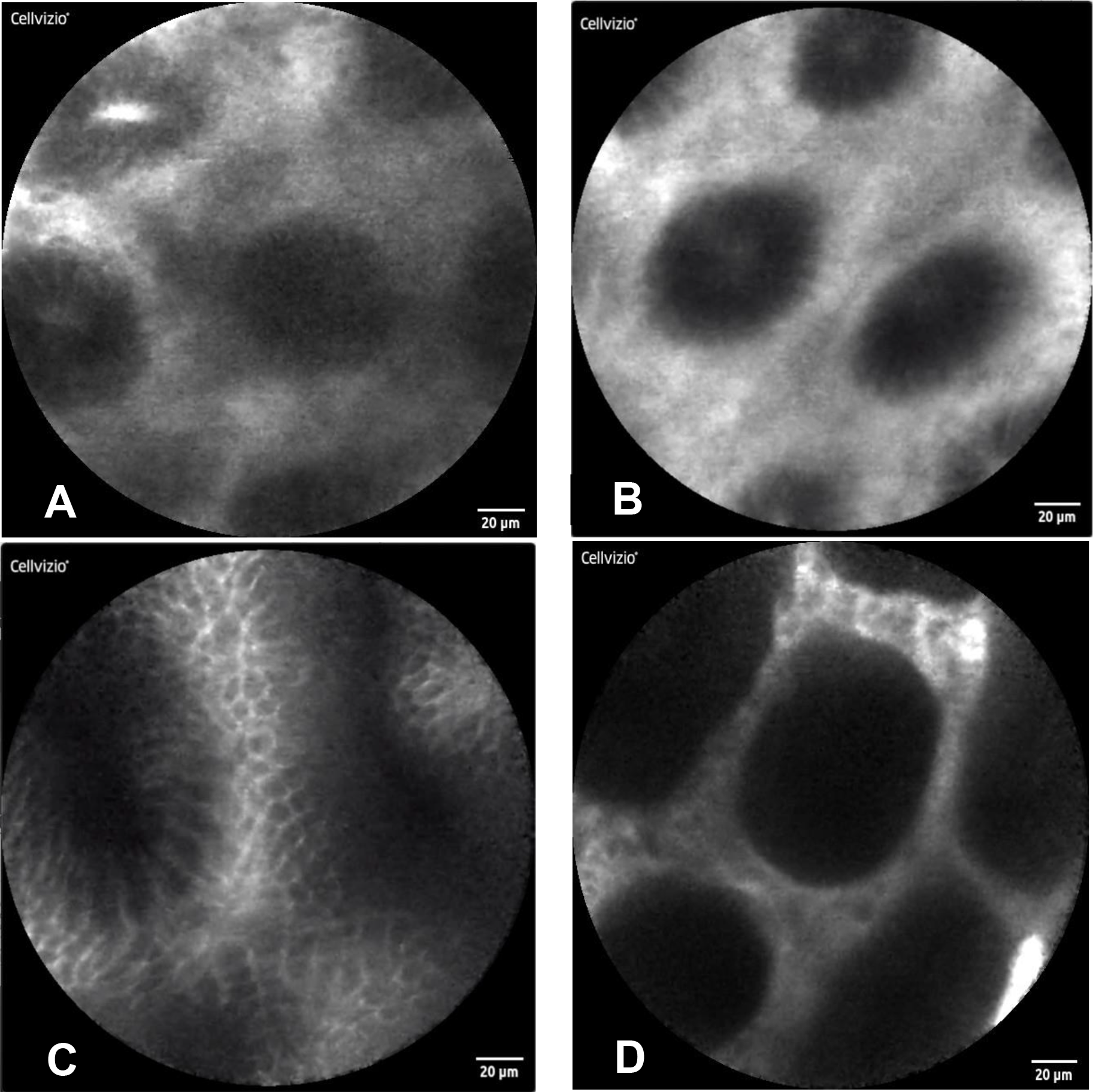
Probe-based CLE images of gastric mucosa interpreted as normal and confirmed by biopsy with histopathology. Normal appearing confocal laser endomicroscopic image of gastric fundus (a), gastric body (b), gastric antrum (c), and gastric incisura (d).

Cambridge method of non-targeted biopsies was performed in all patients following pCLE examination. A total of 1080 CM biopsies were obtained. Occult SRC carcinoma was detected in 4 patients, all from the gastric fundus. The overall detection of occult cancer using CM of non-targeted biopsies was 11.1% (4/36). CM detected SRC carcinoma in 3 of the 6 patients for which pCLE also detected cancer foci. pCLE biopsies detected SRC carcinoma in 3 of the 4 patients for which CM also detected cancer foci. According to surveillance biopsy method, the overall positive biopsy rate was 2.1% (6/282) with pCLE and 0.4% (4/1080) with CM (p< 0.05). At 14 days post-procedure there were no serious adverse events related to pCLE or upper gastrointestinal endoscopy. During one pCLE examination the probe cap detached within the gastric body and was safely retrieved without sequelae.

### True cancer rates

Fifteen patients (41.6%) subsequently elected for prophylactic total gastrectomy. All 15 gastrectomy specimens contained SRC carcinomas (T1aN0). Mean SRC focus size was 1.9mm (range 0.1 – 8.0). In these patients, pCLE successfully detected occult SRC in 5/15 (33.3%) patients, whereas CM detected SRC in 2/15 (13.3%). A comparison of the pCLE and CM detection methods in the 15 patients who underwent prophylactic total gastrectomy revealed the following results: 2 of the 15 patients had SRCs identified by both methods; 10 of the 15 patients did not have SRCs detected by either method; 3 of 15 patients had SRCs that were identified by pCLE but not by CM. There were no instances of SRC being found by CM but not found by pCLE. Thus, 12/15 (80%; 95% CI: 52–96%) were in agreement by both methods. The false negative detection rates for pCLE and CM were 66.7% (10/15) and 86.7% (13/15), respectively. Most SRC foci detected endoscopically were from biopsies obtained from the fundus (84.6%, 11/13). Probe-based CLE biopsies detected an additional 2 foci of SRC in the gastric body, whereas CM biopsies from the body, incisura, and antrum were negative for cancer in all patients (Supplemental Table 1). Upon total gastrectomy explant analysis, foci of cancer were reported in both the cardia/fundus and body, but not at the incisura or antrum. There was no difference in cancer detection rates based on *CDH1* variant type by endoscopic biopsy or total gastrectomy.

## Discussion

In this single-institution phase II study we demonstrated that although pCLE alone may have limited capabilities, it could serve as an adjunct to standard endoscopy to improve occult cancer detection in *CDH1* variant carriers. Probe-based CLE biopsies demonstrated a lower false negative rate of occult cancer detection when compared to both CM biopsy and WLE targeted biopsy of mucosal abnormalities. These results underscore the need for development of techniques that will improve the yield of endoscopic surveillance for early cancer detection in patients with HDGC. Although international consensus guidelines recommend prophylactic total gastrectomy for patients with pathogenic *CDH1* variants, many choose to delay or never pursue this procedure, while others may not be appropriate surgical candidates. Furthermore, some patients decide to pursue gastrectomy only after cancer foci have been identified on endoscopic biopsy. Patients and clinicians alike desire an endoscopic method of reliable early detection, and in some cases monitoring of known cancer foci, yet current surveillance techniques remain insensitive.

Beyond the previously reported retrospective cohort studies and case series, this is the first prospective clinical trial of endoscopic surveillance for cancer in *CDH1* variant carriers.^12, 13, 20, 21^ Our overall rate of occult carcinoma detection in this study was 19%. By comparison, Mi et al reported an overall 61% SRC detection rate in *CDH1* pathogenic variant carriers at an expert center. However, based on initial (single) endoscopy, their reported detection rate was 39%, which is similar to rates of 40–50% described by others.^20, 22^ Even so, the rates of occult gastric cancer detection using the Cambridge method are not consistent across centers. Two recent reports of surveillance endoscopy using CM describe lower cancer detection rates of 14.6% (7/48)^13^ and 12% (3/25)^23^. In our experience, occult SRC detection with endoscopic surveillance of asymptomatic patients has ranged from 15 to 36% (*unpublished data)*.^19^ Even so, the individual cancer detection rates using pCLE (16.7%) or CM (11.1%) alone are still lower than expected. We did not observe any differences in genotype, sex, race, or ethnicity that would explain low detection rates. Moreover, our endoscopy team (TH, CK) on average performs 10 surveillance endoscopies monthly for *CDH1* variant carriers and patients with the hereditary diffuse gastric cancer syndrome.

Adjuncts to WLE have been evaluated in an attempt to improve sensitivity of early gastric cancer detection in this population. For instance, in the robust study from the Cambridge group, WLE examination of mucosal abnormalities was augmented with autofluorescence imaging and narrow-band imaging (NBI), which we did not use in the current study.^12^ NBI in tandem with WLE has proven helpful for delineating pale areas and has a high negative predictive value for abnormal-appearing vascular and mucosal patterns.^24^ However, pale areas were rarely seen in the current study and are known to be non-specific for SRC foci.^24^ This lack of specificity was corroborated by van Dieren et al., who showed that non-elevated pale lesions revealed SRC histology in 17% of biopsies, whereas the majority revealed no histologic change or simply inflammation.^22^ Targeted biopsy of any focal abnormality in their report demonstrated SRC in only 11% of samples. It follows that occult SRC cancers discovered in our study, both by pCLE and CM, were obtained from grossly normal gastric mucosa, without pale spots or protruding lesions, which is consistent with other reports.^20, 22^ Chromoendoscopy may improve sensitivity by illuminating pale white spots of gastric mucosa and other suspicious lesions, however this technique is reportedly unable to detect SRC foci less than 4mm in size.^25^ Notably, in the current study, the typical size of SRC foci detected on endoscopic biopsy was considerably less than 1mm. This fact alone reinforces the purported limitation of chromoendoscopy for detection of submillimeter SRC foci, which are pathognomonic of HDGC. The addition of endoscopic ultrasound (EUS) was also evaluated in a series of asymptomatic *CDH1* variant carriers but did not improve sensitivity.^26^ It is still worth considering that a combination of endoscopic modalities may improve detection rates, even though this was not demonstrated by the addition of pCLE to the Cambridge method.

While we hypothesize that the pCLE pattern of displaced gastric pits may help identify SRC foci, further study is needed to rule out the possibility that pCLE biopsies captured SRC by chance. However, we did not detect any SRC foci by WLE-targeted biopsy alone. This may be due to the focused nature of this study, with fewer biopsies taken of incidental abnormal findings (e.g. atrophic gastric mucosa). Comparison of pCLE images and videos compared with corresponding biopsies, potentially aided by machine learning, could help to further identify endoscopic pCLE patterns indicative of SRC involvement and is an area of active research by the authors. Interestingly, the gastric fundus was the most common site overall for biopsies that revealed SRC foci (11/13, 84.6%), which held true also for the majority of pCLE-based biopsies (7/9, 77.8%). Comparatively, 45.2% of all SRC foci detected by Mi et al were in the fundus or cardia.^12^ Despite these findings, an anatomic predisposition to SRC development within the stomach, or genotype-phenotype correlations of such, has yet to be determined.

Among patients who underwent risk-reducing total gastrectomy, the false negative rate of SRC detection by pCLE was substantially lower than that of the Cambridge method. Probe-based CLE detected more SRC foci than CM in this subset of patients, with an average of twenty-two fewer biopsies taken per patient. Our finding that early stage gastric cancer was detected in 15/15 (100%) gastrectomy specimens is at the extreme of SRC detection at total gastrectomy from other studies, ranging from 87% to 100%.^15, 16, 24, 27, 28^ A recent review reinforced the challenge of false-negative biopsies by reporting that endoscopic surveillance can miss 45–60% of SRC foci ultimately present in gastrectomy specimens.^14^ It is also unclear whether differences in disease penetrance amongst various *CDH1* genotypes has an impact on early cancer detection in this population. The fact that nearly all patients who undergo prophylactic total gastrectomy harbor early gastric carcinomas suggests otherwise.

Clinical trials for this rare cancer syndrome are scarce and therefore warrant critical appraisal and acknowledgment of limitations. This prospective study of individuals harboring a rare genetic variant was conducted over a short time period by a consistent group of investigators, which should lessen the variability in practice that may be more common in retrospective studies of this patient population. A limitation of this study is the single-arm design, which reduced our ability to fully distinguish pCLE from CM, even though an intra-patient comparison of pCLE and CM was performed. A practical criticism is the added endoscopy time for pCLE and thus the potential inability to generalize to standard medical practice. In addition, mucolytics, simethicone, and acid suppressing medications, which may have improved mucosal visibility and pCLE imaging, were not used. We contend that use of a 2T endoscope maximized efficiency and accuracy in obtaining mucosal biopsies with pCLE, however its use may also be described as impractical. Despite these limitations, eligibility criteria were broad, and enrollment occurred contemporaneously with clinical care in an attempt to avoid selection bias. Operator bias was mitigated by performing pCLE prior to CM such that any abnormalities induced by biopsy or hemorrhage would be reduced. While the study was powered purposefully to detect a large, and clinically meaningful difference in cancer detection, a larger study population would have allowed more observations to inform our appraisal of pCLE as a potential tool for gastric cancer surveillance.

## Conclusion

Probe-based CLE performed with white light surveillance endoscopy has a low rate of detection of occult gastric carcinoma in *CDH1* variant carriers. Ideally, the aim of any surveillance method in this population should be the detection of clinically relevant cancer foci that harbor true potential for progression to advanced gastric cancer. However, our ability to detect these early cancers and predict their biologic behavior is lacking. Endoscopic surveillance will continue to provide only a binary outcome that is imperfect at best and unreliable at its worst. Clinical management including endoscopic surveillance as an alternative to surgery should be tempered by this reality. Further exploration of advanced endoscopic techniques, including confocal endomicroscopy perhaps augmented by artificial intelligence, is warranted.

## Data Availability

Reasonable requests for data will be reviewed by the senior author.

---

**Supplemental Table 1:** Endoscopic findings and biopsy results.

|  | While Light Endoscopy |  | Probe-Based CLE |  | Cambridge Method | Gastrectomy Explant |
| --- | --- | --- | --- | --- | --- | --- |
| Patient | Visual Findings | Biopsy Results | Number of Biopsies | SRC Location / Size (mm) | SRC Location / Size (mm) | SRC Location / Size (largest focus, mm) |
| 1 | Mucosal atrophy | n/a | 7 | n/a | Fundus / 1.0 | n/a |
| 2 | Polyps | Chronic gastritis | 7 | n/a | n/a | Fundus / 8.0 |
| 3 | Retained endoclip; multiple small white scars | n/a | 9 | n/a | n/a | Upper fundus / 1.1 |
| 4 | Polyps | Gastric mucosa with occasional dilated glands | 4 | Posterior fundus / 1.7 | Fundus / 0.7 | Fundus / 2.0 |
| 5 | Mucosal atrophy, polyps | n/a | 9 | Posterior fundus / 1.6 | Fundus / 0.5 | Upper fundus / 3.0 |
| 6 | Erythematous and ulcerated antral mucosa; mucosal atrophy | n/a | 8 | n/a | n/a | n/a |
| 7 | Nodule in antrum, erythematous mucosa in body | Antrum type gastric mucosa with mild chronic inflammation | 7 | n/a | n/a | n/a |
| 8 | Erythematous mucosa, body | n/a | 5 | n/a | n/a | n/a |
| 9 | Normal | n/a | 6 | n/a | n/a | n/a |
| 10 | Nodular mucosa in fundus and body, friable mucosa | n/a | 7 | n/a | n/a | Body (greater curvature) / 0.3 |
| 11 | Polyps | n/a | 6 | n/a | n/a | Upper fundus / 0.9 |
| 12 | Single polyp | Fundic gland polyp | 6 | n/a | n/a | Fundus / 2.0 |
| 13 | Erythematous mucosa, single papule | Gastric antral mucosa with no pathological changes | 11 | n/a | n/a | n/a |
| 14 | Normal | n/a | 9 | n/a | n/a | n/a |
| 15 | Single polyp | Fundic gland polyp | 10 | n/a | n/a | n/a |
| 16 | Mucosal atrophy; erythematous mucosa in antrum, single polyp | Oxyntic mucosa with a few dilated glands, possibly a fundic gland polyp | 8 | n/a | n/a | n/a |
| 17 | Erythematous mucosa in antrum, possible angioectasia | n/a | 10 | n/a | n/a | n/a |
| 18 | Erythematous mucosa in antrum, possible angioectasia | n/a | 10 | n/a | n/a | n/a |
| 19 | Friable mucosa | Gastric body mucosa with no specific pathology | 8 | n/a | n/a | n/a |
| 20 | Normal stomach | n/a | 6 | n/a | n/a | Body (lesser curvature) / 0.4 |
| 21 | Submucosal tumor with central umbilication in antrum | Antral mucosa, no pathologic changes | 9 | n/a | n/a | Body (greater curvature) / 0.1 |
| 22 | Multiple hyperplastic polyps, erythematous mucosa in fundus | Gastric oxyntic mucosa with mild chronic gastritis with increased eosinophils | 6 | n/a | n/a | n/a |
| 23 | Polyps | Fundic gland polyp | 11 | n/a | n/a | n/a |
| 24 | Friable, nodular mucosa | Chronic active gastritis (Helicobacter pylori positive by immunostain) | 13 | n/a | n/a | Body / 1.0 |
| 25 | Nodular mucosa in fundus and body; polyps, small white spots | Mild chronic gastritis | 7 | n/a | n/a | n/a |
| 26 | Solitary pale spot in fundus | Oxyntic mucosa with no pathologic changes | 7 | n/a | n/a | n/a |
| 27 | Normal | n/a | 7 | Posterior fundus, anterior upper middle body/ 0.1 and 0.9 | n/a | Upper fundus / 2.0 |
| 28 | Nonbleeding erosive gastropathy | Oxyntic and antral type gastric mucosa with focal chronic inflammation | 7 | Posterior upper middle body, posterior fundus/ 0.5 and 0.2 | Fundus / 0.7 | n/a |
| 29 | Polyps | n/a | 8 | Anterior fundus / 0.4 and 0.7 | n/a | Cardia, fundus (anterior + posterior wall), Body (anterior + posterior wall, lesser + greater curvature) / 4.0 |
| 30 | Normal | n/a | 9 | n/a | n/a | Upper fundus / 0.8 |
| 31 | Single papule in antrum; multiple papules in fundus; erythematous mucosa | Antral mucosa with mild chronic gastritis, oxyntic mucosa with mild chronic gastritis | 1 | n/a | n/a | n/a |
| 32 | Normal | n/a | 12 | n/a | n/a | n/a |
| 33 | Erythematous mucosa in antrum | n/a | 8 | n/a | n/a | n/a |
| 34 | Gastritis, single papule | Gastric body type mucosa with no pathological changes | 6 | Anterior fundus / 0.3 | n/a | Fundus, body / 2.0 |
| 35 | Friable mucosa | Chronic gastritis | 6 | n/a | n/a | Body (greater curvature) / <1.0 |
| 36 | Few papules; erythematous mucosa in antrum; nodular mucosa in body | Gastric mucosa with hyperplastic foveolar gland and reactive changes | 12 | n/a | n/a | n/a |

CLE: confocal laser endomicroscopy; SRC: signet ring cells

**Supplemental Figure 1.**
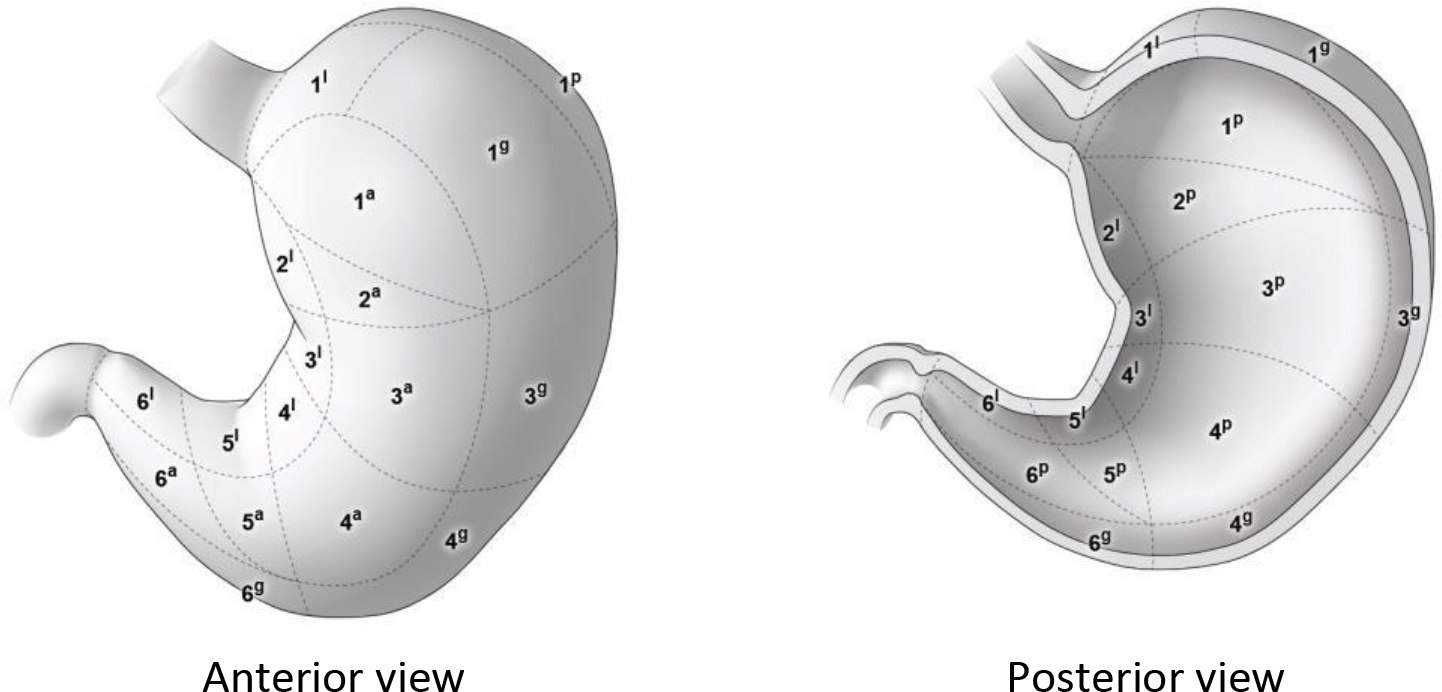
Map of twenty-two anatomic zones for probe-based confocal endomicroscopic evaluation of gastric mucosa. 1, fundus; 2, upper middle body (retroflex); 3, upper middle body (*en face*); 4, lower body; 5, incisura; 6, antrum; a, anterior; p, posterior; l, lesser curve; g, greater curve.

